# Predictors of Death in Severe COVID-19 Patients at Millennium COVID-19 Care Center in Ethiopia: A Case-Control Study

**DOI:** 10.1101/2020.10.07.20205575

**Authors:** Endalkachew H. Maru, Tigist W. Leulseged, Ishmael S. Hassen, Wuletaw C. Zewde, Nigat W. Chamesew, Daniel S. Abebe, Tariku B. Jagema, Abdi B. Bayisa, Mesfin A. Gezahegn, Oli S. Tefera, Wondmagegn G. Shiferaw, Teketel T. Admasu

**Author notes:** Endalkachew H. Maru and Tigist W. Leulseged are joint first authors. Corresponding author: Endalkachew H. Maru, Quality and Clinical Governance Office, Millennium COVID-19 Care Center, Addis Ababa, Ethiopia, Department of Surgery, St. Paul’s Hospital Millennium Medical College, Addis Ababa, Ethiopia.

## Abstract

**Background:** As the number of new cases and death due to COVID-19 is increasing, understanding the characteristics of severe COVID-19 patients and identifying characteristics that lead to death is a key to make an informed decision. In Ethiopia, as of September 27, 2020, a total of 72,700 cases and 1165 deaths were reported.

**Objective:** The study aimed to assess the determinants of death in Severe COVID-19 patients admitted to Millennium COVID-19 Care Center in Ethiopia.

**Methods:** A case-control study of 147 Severe COVID-19 patients (49 deaths and 98 discharged alive cases) was conducted from August to September 2020. A comparison of underlying characteristics between cases (death) and controls (alive) was assessed using a chi-square test and an independent t-test with a p-value of <0.05 considered as having a statistically significant difference. Multivariable binary logistic regression was used to assess a statistically significant association between the predictor variables and outcome of Severe COVID-19 (Alive Vs Death) where Adjusted Odds ratio (AOR), 95% CIs for AOR, and P-values were used for testing significance and interpretation of results.

**Results:** Having diabetes mellitus (AOR= 3.257, 95% CI= 1.348, 7.867, p-value=0.00), fever (AOR=0.328, 95% CI: 0.123, 0.878, p-value= 0.027) and Shortness of breath (AOR= 4.034, 95% CI= 1.481, 10.988, p-value=0.006) were found to be significant predictors of death in Severe COVID-19 patients.

**Conclusions:** The outcome of death in Severe COVID-19 patients is found to be associated with exposures to being diabetic and having SOB at admission. On the other hand, having a fever at admission was associated with a favorable outcome of being discharged alive.

## INTRODUCTION

The 2019 corona virus pandemic has caused significant mortality all over the world, with a total death of 991,000 globally, 25,481 in Africa, and 1165 in Ethiopia as of September 27, 2020. Though it is reported that the African region showed a decreasing trend in the number deaths over the past several weeks compared to other WHO regions, according to the national report the number of deaths in Ethiopia seems considerable over the past weeks, with 10 to 20 death per day, compared to the trend at the beginning of the pandemic in the country ^1,2^.

Studies conducted till today, in a non-African setup, show that the outcome of COVID-19 varies based on individual characteristics and it could include uneventful recovery, respiratory complications requiring invasive mechanical ventilation, renal abnormality requiring kidney replacement therapy, hyper-coagulable state, and death^3-5^.

Studies also identified different risk factors as potential factors for mortality in COVID-19 patients including older age, race, obesity, presence of pre-existing comorbidities, fracture, complications like acute organ failure, lack of early improvement in arterial partial pressure of oxygen (PaO2) to a fraction of inspired oxygen (FiO2) ratio ^4,6-13^.

The role of sex in the severity of the disease or outcome is not clear yet but one study reported no difference in epidemiological characteristics with regard to sex, and in another study, it was reported that sex might have an impact in the clinical presentation and prognosis showing worse prognosis among males ^14-18^.

Abnormal laboratory markers including higher levels of leukocyte count, neutrophil count, highsensitivity C reactive protein, procalcitonin, ferritin, interleukin (IL) 2 receptor, IL-6, IL-8, tumor necrosis factor α, D-dimer, fibrinogen, lactic dehydrogenase, and N-terminal pro-brain natriuretic peptide, raised cytokine and LDH, lower CD4 count and deranged lymphocyte count and others have been attributed as determinants of death in severe COVID 19 patients ^12,18-20^.

Understanding the characteristics of severe COVID-19 patients and identifying those characteristics that lead to death in our setup is the key to make an informed decision to result in the most favorable outcome of treatment for each patient. Yet, there is no such study conducted in Ethiopia.

Therefore, the objective of this study was to assess the predictors of death in Severe COVID-19 patients admitted to Millennium COVID-19 Care Center in Ethiopia.

## METHODS AND MATERIALS

### Study setting and Study population

The study was conducted at the Millennium COVID-19 Care Center (MCCC). The center is the biggest COVID-19 center in the country with a capacity of 1000 bed including ICU patient management.

The study design was a hospital-based case-control study.

The source population was all patients admitted to MCCC with a confirmed diagnosis of COVID-19 using RT-PCR as reported by a laboratory given mandate to test such patients by the Ethiopian Federal Ministry of Health ^21^.

- **Case:** all RT-PCR confirmed COVID-19 infected patients admitted to MCCC and whose treatment outcome was death.
- **Control:** all RT-PCR confirmed COVID-19 infected patients admitted to MCCC and who recovered from the disease and discharged alive.

The study population was all selected COVID-19 patients who were on treatment and follow up at MCCC and who full fill the inclusion criteria.

### Sample Size Determination and Sampling Technique

All COVID-19 related deaths in the Center since the Center started admission on June 2, 2020, up to September 10, 2020 (49 deaths) were included in the study. And with a 1:2 ratio of case to control, 49 deaths and 98 recovered patients, a total of 147 patients were included in the study.

To select the controls, simple random sampling method was used.

### Inclusion and Exclusion Criteria

All COVID-19 patients who were on treatment and follow up at the Center during the observation period were included in the study.

COVID-19 patients who were on treatment and follow at the Center during the observation period but whose outcome status is unknown due to transfer to other hospitals or any other reason that resulted in the discharge of the patient before the observation of the outcome were excluded.

### Operational Definitions

#### Severe COVID-19 disease

Includes patients who have developed complications. The following features can define severe illness ^22^.

∘ Hypoxia: SPO2 ≤ 93% on atmospheric air or PaO2:FiO2 < 300mmHg (SF ratio < 315)
∘ Tachypnea: in respiratory distress or RR>30 breaths/minutes
∘ More than 50% involvement seen on chest imaging

#### COVID-19 Death

A death due to COVID-19 is defined for surveillance purposes as a death resulting from a clinically compatible illness, in a probable or confirmed COVID-19 case, unless there is a clear alternative cause of death that cannot be related to COVID disease (e.g. trauma). There should be no period of complete recovery from COVID-19 between illness and death ^23^.

### Data Collection Procedures and Quality Assurance

Data abstraction tool consisting of all the variables of interest was developed from the patient registration and follow up form and used to abstract the necessary data from the patients’ chart.

Amendment on the tool was made after a pre-test of the data abstraction tool was made on 5% of randomly selected charts.

Training on the basics of the questionnaire and data collection tool was given for four data collectors (General practitioners) and one supervisor (General practitioner).

Data consistency and completeness were checked before an attempt was made to enter the code and analyze the data.

### Data Management and Analysis

The extracted data were coded, entered into Epi-Info version 7.2.1.0, cleaned, stored, and exported to SPSS version 25.0 software for analysis. Categorical covariates were summarized using frequencies and percentages. To assess the presence of a statistically significant difference between those who died and those who were discharged alive in terms of the independent variables, a chi-square test (for the categorical variables), and independent t-test (for the numerical variables) were run. The assumptions of a chi-square test, that no cell should have an expected frequency of less than 5, and the assumptions of t-test, normality of distribution, and equality of variance, were checked before the analysis, and the data was found to satisfy these assumptions. Variables with a p-value of ≤ 0.05 were considered as having a statistically significant difference in terms of COVID-19 outcome (Alive Vs Dead).

The association between the outcome of the disease and predictor variables was assessed using Binary Logistic Regression. Univariate analysis was run at 25% level of significance to screen out independent variables to be used in the Multivariable Binary Logistic regression model. The adequacy of the final model was assessed using the Hosmer and Lemeshow goodness of fit test and the final model fitted the data well (x^2^_(8)_=5.234 and p-value = 0.732). For the Binary Logistic regression, a 95% confidence interval for AOR was calculated and variables with p-value ≤ 0.05 were considered as statistically associated with the outcome of COVID-19 (Alive Vs Dead).

## RESULT

### Socio-demographic, co-morbid illness and medication use related variables and comparison between Alive Vs Dead group

The majorities (25.9%) of the patients were ≥ 70 years and the smallest proportion was in the age range of 50-59 years (15.6%). More than two-thirds (72.8%) of the patients were males. Eighty-seven (59.2%) of the patients had a history of one or more pre-existing co-morbid illness. The commonest being hypertension (33.3%) followed by diabetes mellitus (30.6%), cardiac illness (12.9%), and chronic lung disease (COPD and /or asthma) (11.6%). Twenty patients (13.6%) had a history of medication use including ACEIS, ARBs, and/or NSAIDs within 14 days of admission.

A statistically significant difference in the COVID-19 treatment outcome (Alive Vs Death) was found among the different age groups, those with a history of pre-existing co-morbid illness, hypertension, and diabetes mellitus. Accordingly, a significantly greater proportion of patients in each age group from 27 to 69 years were discharged improved, on the contrary for those ≥ 70 years and above a significant proportion has died (p-value=0.019). Regarding co-morbid illness history, those who died had a significantly higher proportion of patients with one or more pre-existing co-morbid illness (79.6% Vs 20.4%, p-value= 0.0001) compared with those with no co-morbid illness. Similarly, diabetic patients showed a significant association with death outcomes compared to those patients with no diabetes mellitus (53.3% Vs 46.7%, p-value= 0.001). On the other hand, a significant proportion of patients with a history of hypertension (51.0% Vs 49.0%, p-value= 0.004) were discharged alive compared to those with no such illness. (**Table 1**)

**Table 1:**
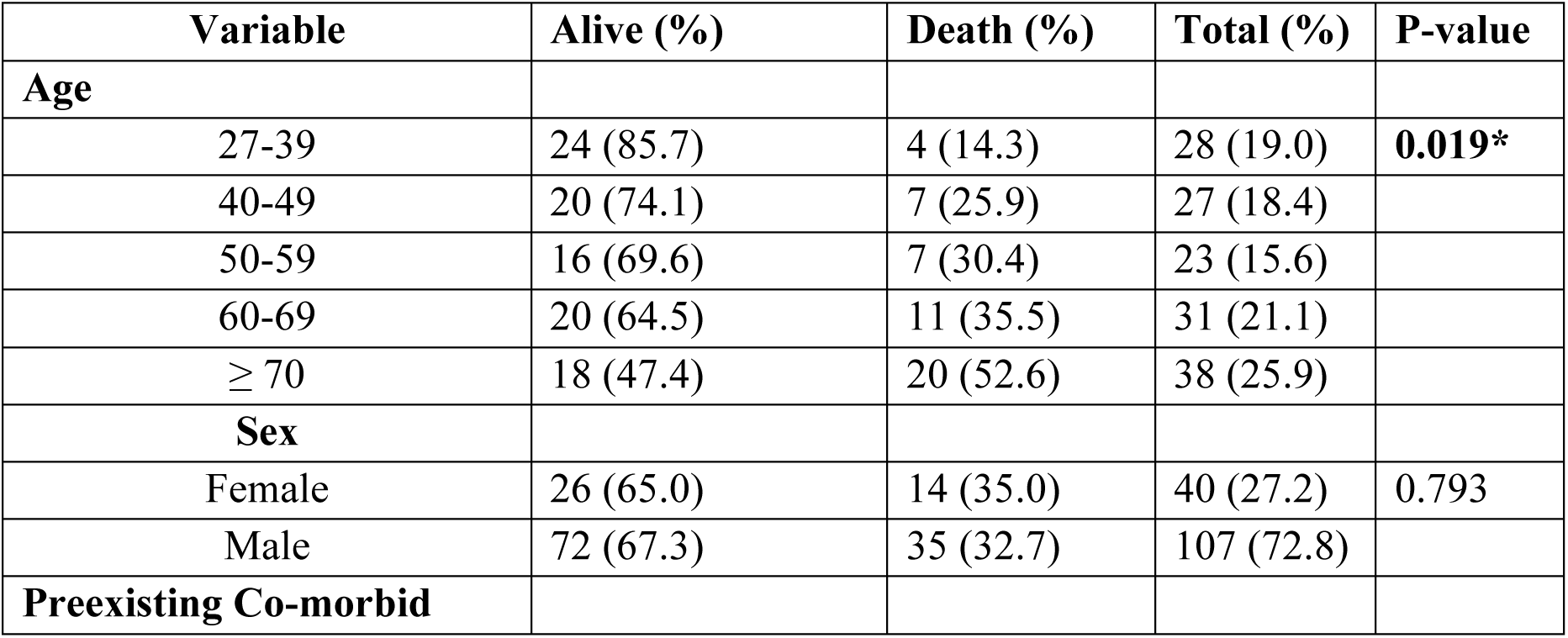
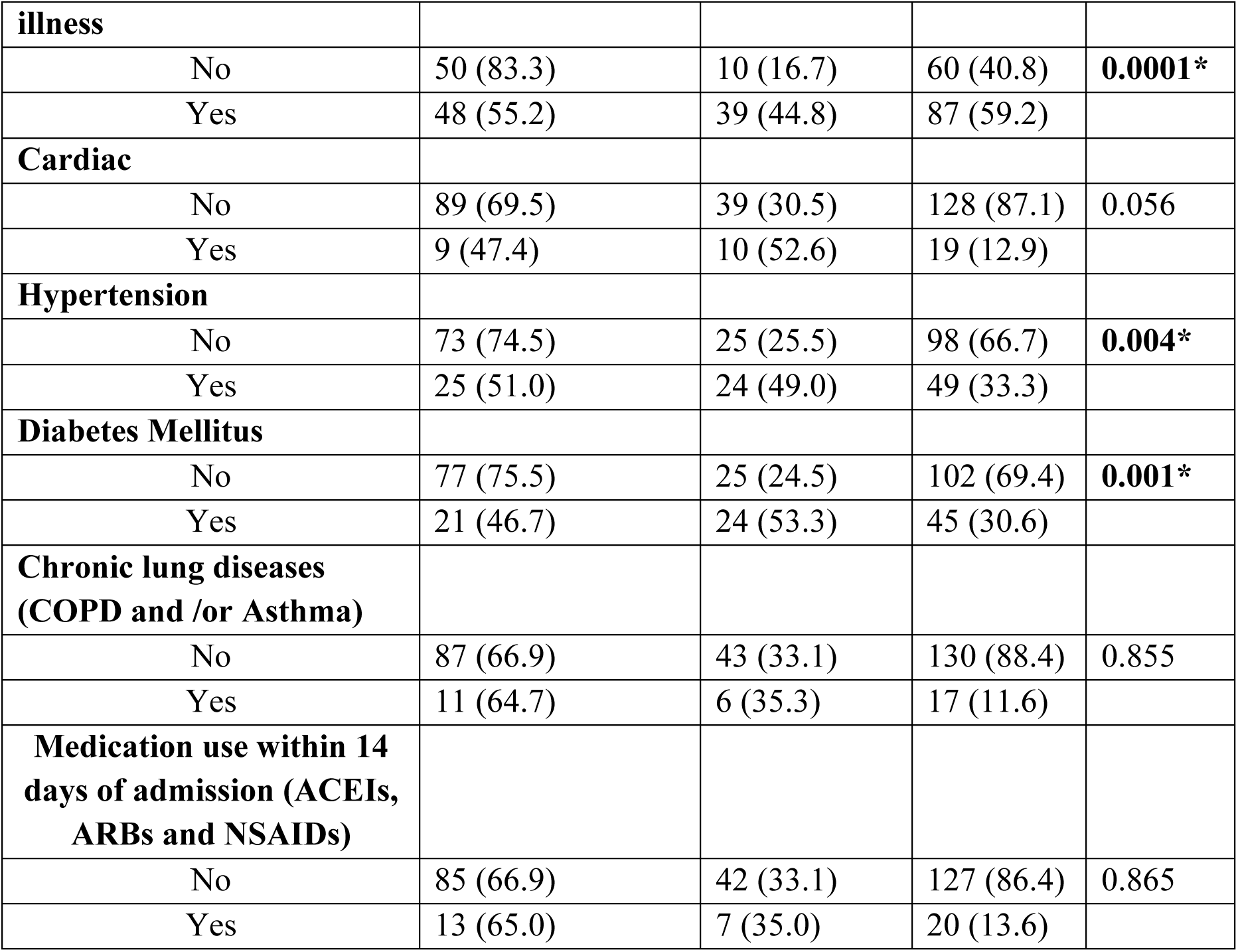
Socio-demographic, preexisting co-morbid illness, and medication use related variables and comparison between Alive Vs dead group among COVID-19 patients (n=147)

### Presenting symptom related variables and comparison between Alive Vs Dead group

Almost all (97.9%) of the patients had a history of one or more symptoms at admission. The majority had cough (82.9%), followed by Shortness of breath (67.3%), fatigue (50.3%) chest pain (36.1%), fever (35.4%), headache (25.9%), sore throat (20.4%), arthralgia (19.1%) and myalgia (17.1%).

According to the chi-square test result, a significantly higher proportion of patients with symptoms of fever (78.8% Vs 21.2%, p-value=0.020), sore throat (86.7% Vs 13.3%, p-value= 0.009) and chest pain (77.4% Vs 22.6%, p-value=0.039) were discharged alive compared to those who had no such symptoms. (**Table 2**)

**Table 2:** Presenting symptom related variables and comparison between Alive Vs dead group among COVID-19 patients (n=147)

| **Variable** | **Controls**<br>**(Discharged**<br>**Alive) (%)** | **Cases**<br>**(Death) (%)** | **Total (%)** | **P-value** |
| --- | --- | --- | --- | --- |
| **Fever** |  |  |  |  |
| No | 57 (60.0) | 38 (40.0) | 95 (64.6) | **0.020\*** |
| Yes | 41 (78.8) | 11 (21.2) | 52 (35.4) |  |
| **Cough** |  |  |  |  |
| No | 15 (60.0) | 10 (40.0) | 25 (17.00) | 0.438 |
| Yes | 83 (68.0) | 39 (32.0) | 122 (82.9) |  |
| **Sore throat** |  |  |  |  |
| No | 72 (61.5) | 45 (38.5) | 117 (79.6) | **0.009\*** |
| Yes | 26 (86.7) | 4 (13.3) | 30 (20.4) |  |
| **Chest pain** |  |  |  |  |
| No | 57 (60.6) | 37 (39.4) | 94 (63.9) | **0.039\*** |
| Yes | 41 (77.4) | 12 (22.6) | 53 (36.1) |  |
| **Myalgia** |  |  |  |  |
| No | 80 (65.6) | 42 (34.4) | 122 (82.9) | 0.535 |
| Yes | 18 (72.0) | 7 (28.0) | 25 (17.1) |  |
| **Arthralgia** |  |  |  |  |
| No | 79 (66.4) | 40 (33.6) | 119 (80.9) | 0.882 |
| Yes | 19 (67.9) | 9 (32.1) | 28 (19.1) |  |
| **Fatigue** |  |  |  |  |
| No | 49 (67.1) | 24 (32.9) | 73 (49.7) | 0.907 |
| Yes | 49 (66.2) | 25 (33.8) | 74 (50.3) |  |
| **SOB** |  |  |  |  |
| No | 37 (77.1) | 11 (22.9) | 48 (32.7) | 0.062 |
| Yes | 61 (61.6) | 38 (38.4) | 99 (67.3) |  |
| **Headache** |  |  |  |  |
| No | 71 (65.1) | 38 (34.9) | 109 (74.1) | 0.505 |
| Yes | 27 (71.1) | 11 (28.9) | 38 (25.9) |  |

### Baseline vital sign and laboratory markers related variables and comparison between Alive Vs Dead group

The independent t-test result shows that, compared to those who were discharged alive, patients who died had a significantly lower mean admission diastolic blood pressure (20.2 mmHg Vs 78.2 mmHg, p-value=0.0001) and mean Spo2 (89.3% Vs 93.1%, p-value=0.001).

On the laboratory biomarkers, those who died had significantly lower mean values of Hemoglobin (13.9 mg/dl Vs 14.9 mg/dl, 0.017, p-value=0.017), lymphocyte percent (4.1% Vs 8.3%, p-value=0.003) and platelet count (245,300 Vs 318,800, p-value=0.002) and higher neutrophil percent (91.8% Vs 82.4%, p-value=0.0001) compared to those who were discharged alive. Similarly, the death group had raised mean values of Urea (66.5 Vs 33.8, p-value=0.0001), creatinine (1.25 Vs 0.82, p-value=0.001) and SGPT (82.0 Vs 47.4, p-value=0.002) compared to those discharged alive. (**Table 3**)

**Table 3:** Baseline vital sign and Laboratory markers related variables and comparison between Alive Vs dead group among COVID-19 patients (n=147)

| **Variable** | **Alive**<br>**(Mean)** | **Death**<br>**(Mean)** | **P-value** |
| --- | --- | --- | --- |
| **Vital sign at presentation** |  |  |  |
| Temperature (°C) | 36.4 | 36.2 | 0.237 |
| Heart rate (Beats/min) | 99.0 | 96.8 | 0.498 |
| Respiratory rate (RR/min) | 26.0 | 27.0 | 0.437 |
| SBP (mmHg) | 131.1 | 125.3 | 0.145 |
| DBP (mmHg) | 78.2 | 20.2 | **0.0001\*** |
| Spo2 (%) | 93.1 | 89.3 | **0.001\*** |
| **Complete cell count** |  |  |  |
| Hemoglobin (mg/dl) | 14.9 | 13.9 | **0.017\*** |
| Hematocrit (%) | 43.5 | 42.5 | 0.411 |
| White blood cell count (Cells/L) | 64.3 | 14.9 | 0.510 |
| Neutrophil % (%) | 82.4 | 91.8 | **0.0001\*** |
| Absolute neutrophil count<br>(cells/L) | 12.2 | 13.8 | 0.616 |
| Lymphocyte % (%) | 8.3 | 4.1 | **0.003\*** |
| Platelet count (cells/ L) | 318.8 | 245.3 | **0.002\*** |
| **Renal function test** |  |  |  |
| Urea (mg/dl) | 33.8 | 66.5 | **0.0001\*** |
| Creatinine (mg/dl) | 0.82 | 1.25 | **0.001\*** |
| **Liver function test** |  |  |  |
| Asparatate transaminase (IU/L) | 60.3 | 67.9 | 0.540 |
| Alanine transaminase (IU/L) | 47.4 | 82.0 | **0.002\*** |
| Alkaline phosphatase (IU/L) | 89.1 | 113.9 | 0.123 |
| **Electrolyte** |  |  |  |
| Sodium (mEq/L) | 149.5 | 139.2 | 0.576 |
| Potassium (mEq/L) | 4.36 | 4.43 | 0.752 |

### Factors associated with death outcome in Severe COVID-19 patients

Based on the result of the univariate analysis at 25% level of significance; Age group, hypertension, diabetes mellitus, fever, sore throat, chest pain and SOB were found to be significantly associated with COVID-19 treatment outcome.

However; only history of diabetes mellitus, fever and SOB were found to be significantly associated with COVID-19 treatment outcome (Alive Vs dead) in the multiple Binary Logistic Regression model at 5% level of significance.

Accordingly, after adjusting for other covariates, the odds of dying among those with a history of diabetes were 3.257 times compared to those with no diabetes (AOR= 3.257, 95% CI= 1.348, 7.867, p-value=0.009).

Patients who died were 67.2% less likely to have a history of fever at admission compared with those who recovered from the disease (AOR=0.328, 95% CI: 0.123, 0.878, p-value= 0.027).

On the other hand, the odds of dying among patients who presented with shortness of breath at admission was 4.034 times compared to those with no such symptom (AOR= 4.034, 95% CI= 1.481, 10.988, p-value=0.006). (**Table 4**)

**Table 4:** Results for the final multivariable binary logistic regression model among COVID-19 patients (n=147)

| Variable | COR (95% CI) | AOR (95% CI) | P-value |
| --- | --- | --- | --- |
| **Age** |  |  |  |
| 27-39 | 1 | 1 | 0.185 |
| 40-49 | 2.100 (0.537, 8.217) | 1.746 (0.383, 7.950) | 0.471 |
| 50-59 | 2.625 (0.659, 10.453) | 1.175 (0.237, 5.816) | 0.843 |
| 60-69 | 3.300 (0.909, 11.977) | 1.449 (0.309, 6.803) | 0.638 |
| ≥ 70 | 6.667 (1.938, 22.929) | 4.445 (0.987, 20.024) | 0.052 |
| **Hypertension** |  |  |  |
| No | 1 | 1 |  |
| Yes | 2.803 (1.363, 5.765) | 1.321 (0.522, 3.347) | 0.557 |
| **Diabetes Mellitus** |  |  |  |
| No | 1 | 1 |  |
| Yes | 3.520 (1.681, 7.372) | **3.257 (1.348, 7.867)** | **0.009\*** |
| **Fever** |  |  |  |
| No | 1 | 1 |  |
| Yes | 0.402 (0.184, 0.880) | **0.328 (0.123, 0.878)** | **0.027\*** |
| **Sore throat** |  |  |  |
| No | 1 | 1 |  |
| Yes | 0.246 (0.081, 0.752) | 0.277 (0.076, 1.008) | 0.051 |
| **Chest pain** |  |  |  |
| No | 1 | 1 |  |
| Yes | 0.451 (0.210, 0.969) | 0.616 (0.244, 1.557) | 0.306 |
| **SOB** |  |  |  |
| No | 1 | 1 |  |
| Yes | 2.095 (0.955, 4.597) | **4.034 (1.481, 10.988)** | **0.006\*** |

**Note:** COR, Crude Odds ratio; AOR, Adjusted Odds ratio; CI, Confidence interval; \*Statistically significant

## DISCUSSION

In this study, we assessed the predictors of death in severe COVID-19 patients who were admitted at MCCC. The chi-square and independent t-test result showed that there was a significant difference in the underlying characteristics of patients who died and discharged alive, showing that the death group was significantly older, had one or more comorbid illness, had diabetes mellitus, had lower diastolic blood pressure and lower SPO_2_, lower hemoglobi, neutrophil predominance, relatively lower platelet count and raised urea, creatinine, and ALT. On the contrary, a significantly lower proportion of patients with hypertension, symptoms of fever, sore throat, and chest pain died compared to those discharged alive. This result shows the possible contribution of socio-demographic and clinical characteristics in disease outcome in the study setup.

On the univariate analysis; Age group, hypertension, diabetes mellitus, fever, sore throat, chest pain, and SOB were found to be significantly associated with COVID-19 treatment outcome showing that these factors significantly contribute to disease outcome independently.

On further analysis after controlling for other covariates, using a multivariable binary logistic regression, only three factors were found to be significant predictors of death in patients with severe disease; a history of diabetes mellitus, fever, and Shortness of breath at admission.

Being diabetic was found to be an important predictor of disease outcome. The odds that those who died had a history of diabetes were 3.257 times compared with those who recovered from the disease. This could be because diabetes mellitus, especially if poorly controlled, is known to lead to compromised immunity that decreases the body’s ability to fight off any infection including viral infections like COVID-19. Also, the chances of having and/or developing another chronic illness are higher than non-diabetic individuals. This results in any diabetic patient to be vulnerable to develop symptomatic infection and complications from any infectious disease that could result in worse disease prognosis. This result also seems to be the case in others’ setup as it is reported that having diabetes mellitus is associated with poor prognosis in studies conducted in China and England ^14,17-19^.

The study found that having fever at admission is associated with having a favorable outcome of being discharged alive. Patients who died were 67.2% less likely to have a history of fever at admission compared with those who recovered from the disease. As fever is an indication of serious disease, it is also an indication of an active immune response of the body to infection including a virus in an attempt to prevent it from overtaking the body. Therefore, having symptoms of fever could imply that the patient has a competent immunity that is fighting the infection which in turn leads to a favorable disease outcome. In other words, a compromised immunity due to different causes means the immune system may not mount the ordinary reactions to threats like fever which leads the person to have a higher risk of serious infection.

In addition, the result of the study found that having SOB at admission is one of the significant factors that predict having a death outcome. The odds that those who died had a history of SOB at presentation were 4.034 times those who recovered from the disease. Shortness of breath is a manifestation of decreased lung function and is considered as a sign of a life-threatening condition. Patients with persistent shortness of breath complaint are those with such lower lung reserve, functional or anatomical cause, that makes them vulnerable to a virus like COVID-19 and are unable to cope with the stress leading to a bad prognosis.

On the other hand, the other risk factors identified in most studies like old age, male sex, and hypertension didn’t show any significant effect on disease outcome in the final regression model ^14,17,18,20^. Age group and hypertension showed a significant association in the chi-square test and a significant association on the univariate binary logistic regression but didn’t show any predictive effect on the multivariable binary logistic regression. On the other hand, Sex didn’t show any association or predictive value on the chi-square test and binary logistic regression model (both univariate and multivariate).

The other important determinants identified in most studies are deranged laboratory parameters ^12,18-20^ but in our study, the only laboratory parameters assessed for all patients are complete blood cell count, renal function test, and liver function, and none showed any significant association. Other relevant parameters were not assessed for the patients during the clinical course due to the unavailability of the laboratory tests and reagents.

## CONCLUSION

Being diabetic and having SOB at admission were found to be predictors of death in Severe COVID-19 patients. On the other hand, having a fever at admission was associated with a favorable outcome of being discharged alive.

We recommend a more careful follow-up and management of diabetic patients. In addition, a more strict infection prevention practice should be advised to diabetic individuals. A more careful evaluation of patients with SOB should be done with a more sensitive triaging method to pick the symptom. Furthermore, SOB can be used as a warning sign in a patient follow-up to provide early detection and intervention for a favorable outcome.

## Data Availability

All relevant data are available upon reasonable request

## Declaration

### Ethics approval and consent to participate

The study was conducted after obtaining ethical clearance from St. Paul’s Hospital Millennium Medical College Institutional Review Board. Written informed consent was obtained from the participants. The study had no risk/negative consequence on those who participated in the study. Medical record numbers were used for data collection and personal identifiers were not used in the research report. Access to the collected information was limited to the principal investigator and confidentiality was maintained throughout the project.

### Authors Contribution

EMH, TWL and ISH conceived and designed the study, revised data extraction sheet and drafted the initial manuscript. ABB and TBJ designed data extraction sheet. All authors contributed to the conception and obtained patient data. EMH and TWL performed statistical analysis. All authors undertook review and interpretation of the data. All authors revised the manuscript and approved the final version.

## Acknowledgment

The authors would like to thank St. Paul’s Hospital Millennium Medical College for facilitating the research work.

## Availability of data and materials

All relevant data are available upon reasonable request.

